# Predicting severity of Covid-19 using standard laboratory parameters

**DOI:** 10.1101/2021.01.07.21249392

**Authors:** Thimo Buchmüller, Ingmar Gröning, Ralf Ihl

## Abstract

**Background:** More than 1.6 million people have already deceased due to a COVID-19 infection making it a major public health concern. A prediction of severe courses can enhance treatment quality and thus lower fatality and morbidity rates. The use of laboratory parameters has recently been established for a prediction. However, laboratory parameters have rarely been used in combination to predict severe outcomes.

**Method:** We used a retrospective case-control design to analyze risk factors derived from laboratory parameters. Patients treated for COVID-19 at an hospital in Krefeld, Germany, from March to May 2020 were included (n =42). Patients were classified into two categories based on their outcome (Mild course vs. treatment in intensive care unit). Laboratory parameters were compared across severity categories using non-parametric statistic. Identified laboratory parameters were used in a logistic regression model. The model was replicated using a) clinical standardized parameters b) aggregated factors derived from a factor analysis.

**Results:** Patients in intensive care unit showed elevated ALT, CRP and LDH levels, a higher leukocyte and neutrophile count, a higher neutrophile ratio and a lowered lymphocyte ratio. We were able to classify 95.1% of all cases correctly (96.6% of mild and 91.7% of severe cases, *p*<.001).

**Conclusion:** A number of routinely collected laboratory parameters is associated with a severe outcome of COVID-19. The combination of these parameters provides a powerful tool in predicting severity and can enhance treatment effectiveness.

---

Starting in December 2019 COVID 19 spread to a pandemic disease. In January 2021 more than 84 million people were infected, and more than 1,8 million deceased ^1^. The time interval between infection and start of symptoms is estimated with a median of 5 days ^2^. Patients are seen as infectious in a phase between 2 days before and about 5 days after developing symptoms. From this point in the course of the disease, a median of 4 days is needed to decide, if hospital care will become necessary. For about 14 percent of patients a few days later, intensive care might be unevitable^3^. An early prediction of critical courses can increase the survival rate, for instance by treating an hyperinflammatory syndrome^4^. Previously identified risk factors for fatal outcomes are male gender, old age and comorbidities like hypertension, heart disease and diabetes type 2^5^. However, a save prediction who of the hospitalized patients will need intensive care remains difficult. In order to enhance the predictive quality an increasing number of studies have investigated predictive features of laboratory parameters. The majority of these studies have reported elevated cytokines, altered hematological parameters, increased coagulation factors and increased liver enzymes in severe cases (see ^6^ for a review). Insights into clinically relevant parameters can enhance the predictive quality, however, a more powerful approach is to combine these biomarkers using regression analysis in order to account for common physiological features in severe cases. There is a general scarcity on studies investigating laboratory markers in covid-19 to predict severe courses. Moreover, studies combing these features have very rarely been conducted on this issue. Here, to estimate the course of the disease, the predictive value of routine blood count was investigated using a) pairwise comparisons on single biomarkers, b) using binary logistic regression models to estimate their combined predictive power. Moreover, factor analysis was used to reduce the number of biomarkers used and thus increase the clinical feasibility.

## Methods

The ethical commission of the medical faculty of the Heinrich Heine University Düsseldorf gave approval to the study design (Nr. 2020-999). We used a retrospective case-control design to assess potential predictors of the course of Covid-19 infections using standard laboratory parameters. All data were anonymized prior data analysis.

### Sample and study design

All patients were treated in hospital due to a COVID-19 infection and treatment had been completed at the time of data analysis. Inclusion criteria for Covid-19 patients were 1) at least one positive PCR-test for Covid-19 and 2) a known endpoint of the disease (cured/death). A total of 42 patients with Covid-19 fulfilled the inclusion criteria and were included in the sample. The average age was 70 years (31-94, SD: 18 yrs.), 17 patients were women (40%).

### Data collection

Raw data were collected from trained medical staff using the local electronical records. Data were double checked for correctness, subsequently anonymized and transferred to the first authors for data analysis. Data were collected from March to May, 2020 at the Maria-Hilf-Hospital of the Alexian Krefeld GmbH, a multidisciplinary hospital in Germany. To guarantee the anonymity of patients we only included age and gender as demographic characteristics.

### Laboratory parameters and clinical data

Since laboratory parameters were available at several points in time, the parameters closest to the sampling for Covid-19 were used. Laboratory parameters were based on the standard assessments regularly conducted for elderly patients. A total of 11 parameters were included, which can be classified into ‘biochemical chemistry’ (ALT), ‘inflammation markers’ (CRP, Urea), ‘myocardial enzymes’ (LDH), ‘coagulation factors’ (Quick, PTT) and ‘hematology’ (Neutrophils (% and count), lymphocytes (% and count) and leukocytes (count)). Clinical characteristics for Covid-19 patients were firstly survival (cured/died), and treatment in ICU or palliative treatment, respectively. Of the 42 patients 9 were treated in ICU and further 4 received palliative care. Due to the relatively low number of cases which were treated in palliative care we aggregated the end-points ‘palliative care and ‘treated in ICU’ obtaining a ‘severe infection’ (1) and compared it to ‘mild’ cases, namely covid-19 patients which were not treated in ICU.

### Data preparation and statistical analysis

In order to estimate the predictive power of laboratory parameters we used a three-step approach. First, pairwise comparisons (Mann-Whitney U test) were performed on all laboratory parameters using infection severity (mild vs. severe) as independent factor (*p*_*exact*_<.05, two-sided). To adjust for multiple comparisons we used Bonferroni-Holm correction. As many parameters (ALT, CRP, LDH, lymphocytes count, Quick, PTT) deviated significantly from normality (Kolmogorov-Smirnov test *p*_exact_<.05) we used non-parametric statistics which requires fewer assumptions. Missing cases were deleted pairwise. Subsequently, all laboratory values which differed between the two groups at least on trend level (p<.1) were used in a binary logistic regression model to predict infection severity (mild (0) vs. severe cases (1)). Nagelkerke’s R square was used to assess the overall model quality (0.2 →acceptable, 0.4→ good, 0.5→ very good). We estimated the parameters using bootstrapping (BcA, n = 500, k = 20000), which is more robust to outliers and allows to simulate a larger sample size. Missing cases were deleted listwise, if more than 2 out of 6 parameters were missing. If fewer parameters were missing we used the mean of the time series (1 case). The logistic regression was repeated using the same laboratory parameters standardized to their clinical significance. Here, parameters (our predictors) were classified in a binary way as 0 indicating that patients were within a normal range or as 1 if parameters were beyond a normal range, indicating a potential clinical significance. Norm values were used based on standardized laboratory conventions. In order to develop a risk score obtained from the previously identified parameters we conducted further analysis steps. Using a factor analysis (varimax rotation) on the previously used laboratory parameters we aimed at aggregating biomarkers. The Kaiser-Gutmann criteria was used to limit the number of extracted factors. Subsequently, a risk score was calculated using the obtained factor structure. Here, laboratory parameters were added up within the factor they loaded highest on and divided by the number of parameters of the respective factor. The clinical standardized laboratory parameters were used here in order to enhance the feasibility of the risk score for clinical use. Analysis were performed with SPSS version 26. (IBM).

## Results

### Pairwise comparisons

Most laboratory parameters differed across infection groups (see Tab.1). Specifically, we found elevated ALT, CRP and LDH levels in severe cases as well as a higher leukocyte and neutrophile count and a higher neutrophile ratio in severe cases, while the lymphocyte ratio was lowered in severe cases. The coagulation factors (PTT, Quick), urea and the lymphocyte blood count did not differ across outcomes.

### Predicting severity of corona

We combined the previously identified parameters using a binary logistic regression model to predict infection severity. However, we only used hematological blood ratio for neutrophiles and lymphocytes, respectively, due to their high collinearity with the blood count parameter (Spearman-rank correlation: Lymphocytes count*ratio r = .64, neutrophile count*ratio r = .69, all *p*<.001). Average age and gender distribution were similar in both outcome groups (Age: Mann-Whitney U *p*_exact_=.49, Gender: Chi-Square *p*_exact_=.53) and were thus not entered as covariates.

Using six parameters as predictors for the severity of an infection we were able to classify 95.1% of all cases correctly (Nagelkerke’s R = .80, chi-square = 33.65, *p*<.001), which indicates a very good model fit. Specifically, 96.6% of mild cases, respectively, 91.7% of severe cases were classified correctly. Three parameters allow a significant independent contribution in our model, namely CRP, and the lymphocyte and neutrophile ratio (*p*<.05). In order to enhance the clinical feasibility, we repeated the binary regression model entering clinical standardized laboratory predictors (categorical predictors→ higher/lower than norm)). Here, the accuracy was markedly lower and dropped to 88%. The model fit was still good (Nagelkerke’s R = .44, chi-square = 14.5, *p*<.05), however, the number of correctly classified severe cases was low (64%), whereas the model worked well in predicting mild cases (97%).

### Factor analysis and risk score development

A principal component analysis was conducted on the obtained six laboratory parameters. As the factor structure differed markedly between patients treated in ICU (3 factors) and those not treated in ICU (2 factors) we did not perform the factor analysis across all patients. Due to a better estimated sampling adequacy and better sphericity indicated by the Kaiser-Meyer-Olkin (KMO) criteria, respectively, the Bartlett’s test for sphericity (BTS) we selected only patients not treated in ICU for our subsequent factor analysis. Here, the KMO verified our sampling adequacy (KMO = .63) and BTS was *χ*^*2*^ (15) = 176.47, *p* < .001, indicating that the correlations between the items were large enough for the factor analysis. Two factors were extracted with an eigenvalue greater 1 explaining 77% of the total variance. Leukocytes, lymphocytes and neutrophiles loaded high on factor 1, whereas CRP and LDH loaded high on factor 2.

We calculated a risk score derived from the parameters loading highest on a specific factor. These two factors were subsequently entered in a binary regression model. Our two factors predicted a treatment in ICU significantly (p<.05), however with only acceptable model fit (Nagelkerke’s R square = .24). A total of 77% cases were classified correctly, however the risk score performed poorly in predicting those patients with a severe infection (40% correct).

## Discussion

A small number of laboratory values, which are routinely collected, allow a prediction of whether patients have a severe course of a COVID-19 infection with a probability of 95 percent. These parameters could be used in routine care for an evidence-based estimation of the future course and be used along with other critical values to adapt the treatment early in the diagnostic process, thus enhancing treatment quality. Our results are in line with previous studies reporting altered laboratory parameters in severe cases. For instance, CRP and LDH were elevated in COVID-19 patients in general^7^ and also higher in severe cases^8,9^. Similarly, an elevated count of neutrophile and leukocytes are in line with early clinical reports^10^. Interestingly, coagulation factors and lymphocytes did not differ between outcome groups in our study, which is at odds with previous studies^6^. Partly this could be explained by different comparisons groups as coagulation factors were mostly elevated in patients with fatal outcome ^11^. A post-hoc analysis in our sample showed a trend towards increasing PTT values during an infection as compared to PTT before the infection, however only in small subsample (7 patients with available laboratory data before their COVID-19 infection, not reported here). Thus, a larger sample with more cases with fatal outcome might yield similar results. While our study mostly replicated previous findings with regard to single laboratory parameters, only very few studies showed how they work in combination to predict severe courses. The – to our knowledge-only study using a logistic regression model found independent effects of neutrophile and leukocyte with large effect sizes in predicting fatal outcomes^12^. In line with this study we found independent effects of neutrophiles and leukocytes, however, we added CRP as an independent risk factor. Moreover, the authors concentrated on the independent effects of single risk factors, whereas our main aim was to test their combined power. Furthermore, they used categorical parameters with defined clinical thresholds, whereas we showed that widely used clinical cutoffs decrease the statistical power.

Our analysis revealed two limitations regarding the clinical utility of previous approaches. Firstly, widely used clinical thresholds are not well suited to predict the course of COVID-19. This stresses the need to adapt the thresholds to COVID-19 patients to enhance their clinical utility. A second caveat can be derived from the results of our factor analysis. Here, factor structures of the laboratory parameters differed between patients with mild as compared to those with severe infection courses (although a larger sample is needed to replicate the factor structure in severe cases). Hence, researchers developing a clinical ‘risk score’ derived from laboratory parameters and demographic variables might need to consider differential clinical profiles depending on infection severity.

Given our relatively small sample size our results should be interpreted with care. Moreover, one should bear in mind that our findings were based on cross-sectional data and thus point towards laboratory abnormalities in acutely infected severe cases. Here, longitudinal studies are needed to predict future outcomes using the obtained parameters at the beginning of an infection. The differences between predictive quality of raw values as compared to binary values obtained from clinical thresholds and the difference in factor structures points towards the need of estimating a potential course of infection by assessing the combined laboratory profile rather than on defined deviations on specific parameters.

Future research needs to replicate the obtained findings in a larger sample. Moreover, in order to enhance the clinical utility of the obtained findings clinical thresholds for the parameters need to be adapted to patients with severe outcomes. Subsequently, a factor analysis could be used to develop a risk score adapted to patients with a severe course to enhance its predictive power.

Taken together, our study points towards the utility of biomarkers in predicting a severe course of COVID-19, which are widely assessed and can directly be used in routine care. The combination of these parameters is a powerful tool to predict severe outcomes. Along with a range of other known risk factors medical staff can use the obtained parameters to adapt their treatment strategy.

**Table 1.**

| Laboratory parameters | Median (IQR)<br>Non-ICU | Median (IQR)<br>ICU | Mann-Whitney-U, p-value | Effect size g |
| --- | --- | --- | --- | --- |
| ALT | 24.0 | 39.5 | 103.5, .21 | -0.26 |
| CRP* | 3.5 | 14.5 | 53.0, .001 | -1.93 |
| Urea | 36.0 | 81.0 | 94.0, .57 | -0.57 |
| LDH | 217.0 | 346.0 | 85.5, .06 | -1.31 |
| Quick | 86.7 | 83.0 | 173.5, .99 | -0.05 |
| PTT | 25.7 | 24.7 | 155.5, .99 | .21 |
| Neutrophils (%)* | 65.8 | 80.0 | 61.0, .02 | -1.10 |
| Neutrophils<br>(count) * | 3.7 | 6.5 | 54.6, .02 | -1.18 |
| Lymphocytes<br>(%)* | 20.0 | 9.0 | 64.5, .03 | 0.81 |
| Lymphocytes<br>(count) | 1.1 | 0.7 | 99.0, .43 | -0.38 |
| Leukocytes<br>(count) | 5.7 | 7.9 | 112.5, .06 | -.81 |

\*p<.05, two-sided (exact), Bonferroni-Holm corrected
ICU: Intensive Care Unit, corrected effect size hedge's g, IQR: Interquartile range

## Data Availability

Data can be provided upon request.

## References

1. World Health Organization. WHO Coronavirus Disease (COVID-19), Dashboard, 2021. (https://covid19.who.int/).

2. Robert-Koch-Institut. Coronavirus SARS-CoV-2 - und COVID-19, Epidemiologischer Steckbrief zu SARS-CoV-2, 2021. (<https://www.rki.de/DE/Content/InfAZ/N/Neuartiges_Coronavirus/Steckbrief.html;jsessionid=EDB1262CB53A6EA46770CFF9EF22359F.internet121?nn=13490888).

3. Richardson S, Hirsch JS, Narasimhan M, et al. Presenting Characteristics, Comorbidities, and Outcomes Among 5700 Patients Hospitalized With COVID-19 in the New York City Area. JAMA 2020;323:2052–9.

4. Mehta P, McAuley DF, Brown M, Sanchez E, Tattersall RS, Manson JJ. COVID-19: consider cytokine storm syndromes and immunosuppression. Lancet 2020;395:1033–4.

5. Grasselli G, Greco M, Zanella A, et al. Risk Factors Associated With Mortality Among Patients With COVID-19 in Intensive Care Units in Lombardy, Italy. JAMA Intern Med 2020;180:1345–55.

6. Velavan TP, Meyer CG. Mild versus severe COVID-19: Laboratory markers. Int J Infect Dis 2020;95:304–7.

7. Li L-Q, Huang T, Wang Y-Q, et al. COVID-19 patients’ clinical characteristics, discharge rate, and fatality rate of meta-analysis. Journal of Medical Virology 2020;92:577–83.

8. Gao Y, Li T, Han M, et al. Diagnostic utility of clinical laboratory data determinations for patients with the severe COVID-19. Journal of Medical Virology 2020;92:791–6. (https://onlinelibrary.wiley.com/doi/pdf/10.1002/jmv.25770?casa_token=JRhTDBx3F-8AAAAA%3A_AadGijAjgFnCghURFgVS0GWnUPGrpyO0TxN6pjX6E9KfnWKoeXDewEqpbAGTTxdApiAPT_QqISfaOoa).

9. Mo P, Xing Y, Xiao Y, et al. Clinical characteristics of refractory COVID-19 pneumonia in Wuhan, China. Clin Infect Dis 2020.

10. Huang C, Wang Y, Li X, et al. Clinical features of patients infected with 2019 novel coronavirus in Wuhan, China. Lancet 2020;395:497–506. (http://www.sciencedirect.com/science/article/pii/S0140673620301835).

11. Tang N, Li D, Wang X, Sun Z. Abnormal coagulation parameters are associated with poor prognosis in patients with novel coronavirus pneumonia. J Thromb Haemost 2020;18:844–7.

12. Hu L, Chen S, Fu Y, et al. Risk Factors Associated With Clinical Outcomes in 323 Coronavirus Disease 2019 (COVID-19) Hospitalized Patients in Wuhan, China. Clin Infect Dis 2020;71:2089–98.

